# Audiometric profiles and patterns of benefit. A data-driven analysis of subjective hearing difficulties and handicaps

**DOI:** 10.1101/2020.04.20.20045690

**Authors:** Raul Sanchez-Lopez, Torsten Dau, William M. Whitmer

## Abstract

**Objective:** Hearing rehabilitation attempts to compensate for auditory dysfunction, reduce hearing difficulties and minimize participation restrictions that can lead to social isolation. However, there is no systematic approach to assess the quality of the intervention at an individual level that might help to evaluate the need of further hearing rehabilitation in the hearing care clinic.

**Design:** A data-driven analysis on subjective data reflecting hearing disabilities and handicap was chosen to explore “benefit patterns” as a result of rehabilitation in different audiometric groups. The method was based on: 1) Dimensionality reduction; 2) Stratification; 3) Archetypal analysis; 4) Clustering; and 5) Item importance estimation.

**Study sample:** 572 hearing-aid users completed questionnaires of hearing difficulties (speech, spatial and qualities hearing scale; SSQ) and hearing handicap (HHQ).

**Results:** The data-driven approach revealed four benefit profiles that were different for each audiometric group. The groups with low degree of high-frequency hearing loss (HLHF) showed a priority for rehabilitating hearing handicaps, whereas the groups with HLHF > 50 dB HL showed a priority for improvements in speech understanding.

**Conclusions:** The patterns of benefit and the stratification approach might guide the clinical intervention strategy and improve the efficacy and quality of service in the hearing care clinic.

## Introduction

The consequences of hearing loss entail activity limitations and participation restrictions (Simeonsson, 2000). The hearing rehabilitation process aims to minimize both aspects and involves two main steps: diagnosis and remediation (Boothroyd, 2007; Goldstein & Stephens, 1981). After the diagnosis of a hearing loss, an intervention strategy that involves a hearing solution, such as a hearing aid (HA), is commonly proposed and selected by the clinician. The compensation strategy chosen in the HA fitting process is, to a large degree, based on the shape of the pure-tone audiogram. The intervention is verified and validated to ensure the quality of the device and the service (Jorgensen, 2016). However, hearing rehabilitation often requires further interaction, such as follow-up visits to adjust the device(s) to the particular needs of the patient, as well as counseling, focused on communication programs and professional advice (Laplante-Lévesque et al., 2010). Overall, each of the steps of the intervention (diagnosis, adjustment and verification) is influenced by technical, personal and social factors (Vestergaard Knudsen et al., 2010).

The evaluation of the efficacy of the hearing rehabilitation process is typically assessed by questionnaires as outcome measures. The questionnaires can be designed to evaluate the individual benefit, the clinical practice or the inclusion of a new device or strategy (Cox et al., 2000; Cox, 2003). Some outcome measures include specific items related to benefit or satisfaction (SADL: Cox & Alexander, 1999; APHAB: Cox & Alexander, 1995; IOI-HA: Cox, 2003; GHABP: Gatehouse, 1999), whereas others are focused on hearing disabilities and handicaps (de Ronde-Brons et al., 2019; SSQ: Gatehouse & Noble, 2004; Hallberg, 1998; Newman et al., 1990). All of these outcome measures aim to capture overall experiences with the hearing rehabilitation (e.g., IOI-HA) and/or specific aspects, such as the speech, spatial and qualities hearing scale (SSQ; Gatehouse & Noble, 2004). The SSQ reflects HA listeners’ current difficulties with respect to speech perception, spatial sound perception and qualities of hearing, such as the ability to follow a conversation, to localize a sound source or to identify a sound. Although the assessment of hearing disabilities is crucial for a successful hearing rehabilitation, the overall benefit does not only depend on auditory disabilities but also on handicaps experienced by the listener (Whitmer et al., 2016), such as the effects on social participation derived from the hearing loss.

One of the primary aims of outcome measures is to quantify the effectiveness of hearing rehabilitation. However, there is no known systematic method to use an individual’s outcome-measure responses to guide any further personalized intervention. The ability of the HCP to understand each patient’s needs and to implement a suitable intervention is, however, crucial for successful rehabilitation (Boothroyd, 2007). In the present study, a data-driven approach, aiming at identifying patterns of benefit in the self-reported outcome measures, was used for exploring the possibilities of personalized interventions. These interventions may be based on the classification of the intervention in “benefit profiles” (optimal, near-optimal, suboptimal) and the important factors that can be addressed to improve the hearing benefit. Furthermore, the auditory deficits associated with different audiometric shapes, were included in such approach by using audiometric-based auditory profiling.

### Stratification based on auditory deficits

The characterization of the hearing deficits of a person in terms of pure-tone audiometry does not typically capture the person’s performance in real-life situations. Therefore, information about supra-threshold auditory deficits, such as speech intelligibility in noise, might help to better understand the scale and scope of an individual’s sensory impairments. Recently, Sanchez-Lopez et al. (2020) proposed a stratification of hearing-impaired individuals into four clinically relevant subgroups, referred to as “auditory profiles.” The auditory profiles were the result of a data-driven analysis that allowed the identification of four archetypal patterns of perceptual deficits along two largely independent dimensions, speech-intelligibility deficits and loudness perception deficits, respectively. Since each auditory profile showed different degrees of deficits, listeners associated with a given profile are likely to experience similar distinct hearing disabilities.

The audiometric thresholds associated with the different auditory profiles showed significant differences. This suggests that different audiometric configurations can be associated with specific supra-threshold deficits. While supra-threshold measures are important, they are not systematically included in audiological assessments. Some situations may limit the amount of time-intensive testing of supra-threshold deficits possible in clinical settings whereas the pure-tone audiometry is currently a required procedure. Therefore, an audiometry-based stratification into “audiometric profiles” or audiometric groups might provide an initial classification and predict the perceptual deficits of the listener. This approach can be useful when applying the stratification to studies without supra-threshold measures retrospectively. However, this pre-classification in audiometric groups does not guarantee that the listener is correctly classified and supra-threshold tests can confirm the listener’s auditory profile (Sanchez-Lopez et al., 2020).

### Aims of the study

In the present study, subjective data from a questionnaire of hearing difficulties (SSQ) and a questionnaire of hearing handicap (HHQ) were analyzed using a data-driven approach and following the principles of the “knowledge discovery from databases” (KDD; Frawley et al., 1992; Mellor et al., 2018). A data set of a clinical population of hearing-aid users was analyzed in an attempt to uncover archetypal “benefit profiles” reflected in the subjective data that can be associated to specific audiometric groups based on the average audiometric thresholds of the four auditory profiles according to Sanchez-Lopez et al. (2020). The goals of the study were; 1) to identify the patterns of benefit associated with different audiometric groups, 2) to identify the priorities of hearing rehabilitation in terms of particular hearing difficulties and handicaps that need to be improved, and 3) to examine the possibilities of implementing a personalized post-fitting rehabilitation strategy.

## Method

The data analysis consisted of five steps, as shown in Figure 1. First, the data (described in the next section) were transformed using factor analysis. Second, the participants were stratified into four groups based on their degree of low-(HL_LF_) and high-frequency (HL_HF_) hearing loss as an approximation of the audiometric profiles of Sanchez-Lopez et al. (2020). Third, to identify archetypal patterns of benefit, the overall data as well as the data belonging to each of the four subgroups were processed using an archetypal analysis. Fourth, the participants were identified as belonging to a cluster of participants showing a similar “benefit profile”, based on their similarity to the archetypal patterns of benefit. Finally, the identified benefit profiles were predicted using supervised learning and the importance of individual questionnaire items was analyzed.

**Figure 1.**
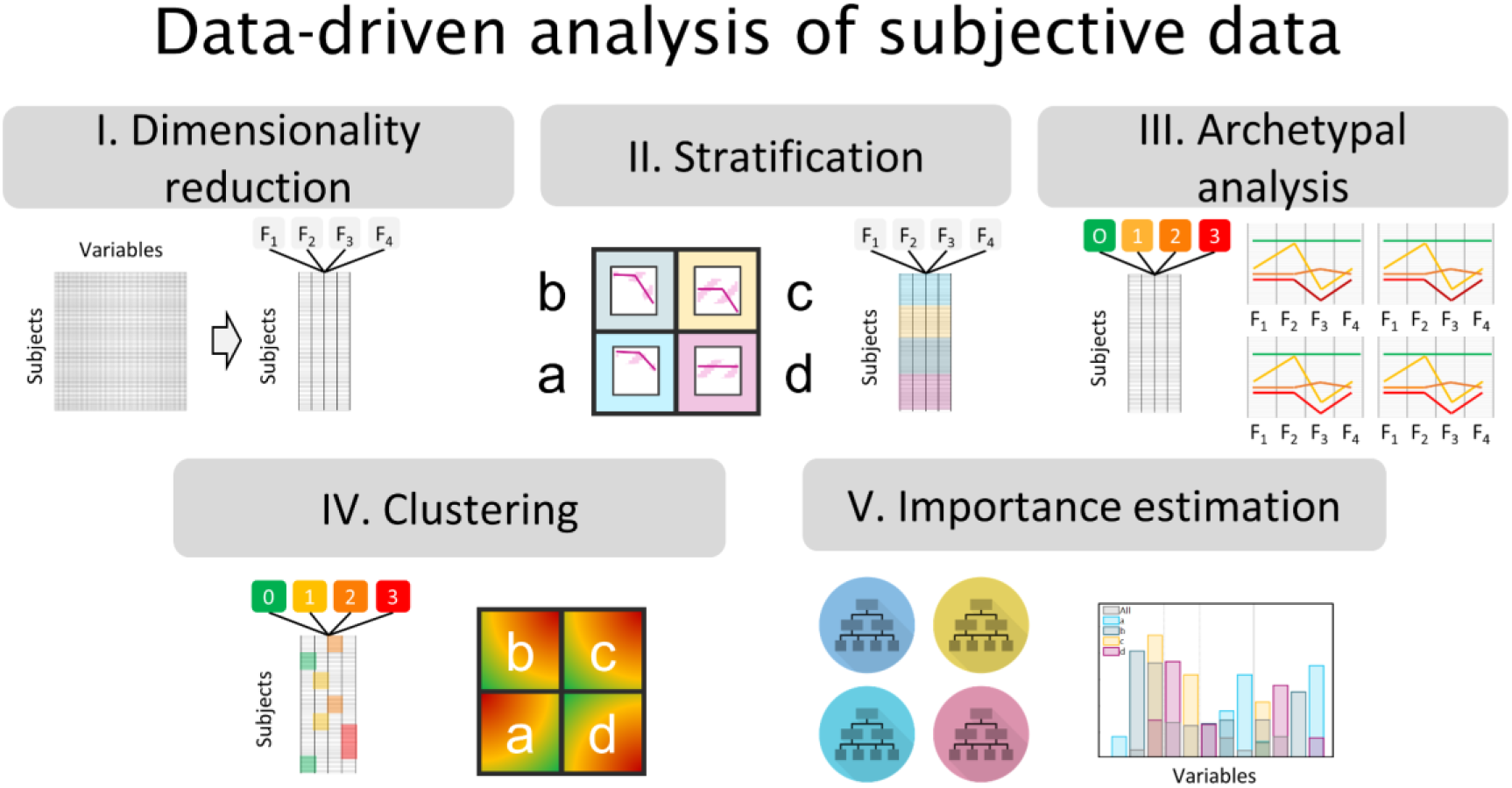
Sketch of the data-driven method for the analysis of the subjective data. Top panel: The unsupervised learning exploratory stages included: I) Dimensionality reduction into four factors (F_1_, F_2_, F_3_, F_4_); II) Stratification, where the subjects were divided into audiometric groups (a, b, c, d); III) Archetypal analysis where the data were decomposed into a matrix with the archetypal patterns of benefit [APoB0, APoB1, APoB2 and APoB3] and the weights of each pattern resembling each subject’s observation. Bottom panel: IV) Clustering, where the participants were clustered based on the similarity of their scores with the benefit profiles derived from stage II; and V) Importance estimation, where a random forest was trained with to classify the participants into the “benefit profiles” and the importance of the predictors were estimated.

### Description of the dataset

The data-driven analysis presented here is a retrospective study performed on a large dataset from Hearing Science Scottish Section (HSSS; formerly Institute of Hearing Research) of the University of Nottingham. The data were collected between the years 2002-2011, and most of the patients were referred from the NHS Audiology clinic of the Glasgow Royal Infirmary. The total dataset consisted of 1471 participants. This HSSS dataset has been previously analyzed by Akeroyd et al. (2014) and Whitmer et al. (2014), where a thorough description of the normative data was provided. The variables of interest for the present study were (1) audiometric thresholds, (2) raw responses to two questionnaires: the Speech, Spatial and Qualities hearing scale (SSQ) and the Hearing Handicap Questionnaire (HHQ; Gatehouse & Noble, 2004). The subset of the HSSS dataset used here consisted of 880 observations (participants), and 62 variables.

### Audiometric thresholds

All thresholds were obtained by trained researchers using the modified Hughson-Westlake adaptive method. Since the data-analysis involved stratification of the participants in audiometry-based auditory profiles, the audiometric thresholds were also retrieved from the dataset but not used in the analysis. The audiometric thresholds were grouped into low-frequency (≤ 1 kHz; LF) and high-frequency (> 1 kHz; HF) averages and only the better ear was used for further analysis. Sanchez-Lopez et al. (2020) did not include participants with average low-frequency hearing thresholds above 65 dB hearing level (HL_LF_) in their data-driven profiling approach. Therefore, to increase the comparability across both stratification strategies, data from participants with a severe-to-profound low-frequency hearing loss (HL_LF_>65dB) were excluded.

### Questionnaire responses

The SSQ consists of 14 speech-understanding (SU) related items, 17 spatial-hearing (SP) related items and 18 qualities-of-hearing (QH) related items. The SSQ questionnaire is scored on a 0-10 scale (in steps of 0.5 in this particular dataset), whereby a low score corresponds to high difficulty and a high score corresponds to low difficulty. If the item corresponds to a situation that the participant has not experienced, the response *not applicable* can be chosen. The 12 HHQ items were based on the Hearing, Disabilities and Handicaps Scale (Hétu et al., 1994). The HHQ is scored on a discrete 1-5 scale, with 5 representing the greatest handicap. Its questions can be divided into two subscales: emotional distress (ED) and social restriction (SR) (Noble et al., 2008). The HHQ data were multiplied by −1, such that a higher value corresponded to a lower handicap, consistent with the scale considered in the SSQ data. Data from participants with 30 or more missing responses across both questionnaires (i.e., missing at least half of the data) were also excluded. All questionnaire data were standardized prior to analysis.

The final number of observations considered for the current analysis was 572 participants (272 female) who wore at least one hearing aid. At the time of testing, 478 (84%) were unilaterally fit (244 left ear, 234 right), the remaining 94 were bilaterally fit. All hearing aids were behind-the-ear from a variety of manufacturers. Median hearing aid experience was 3 years (range 2 wks – 63 years). Their median age was 65 (range 18-85); 71% were ≥ 60 years of age. Their median better-ear four-frequency average was 46 dB HL (range 5-111). The distribution of observations across the four audiometric profiles *a, b, c* and *d* of Sanchez-Lopez et al. (2020) was 63, 81, 345 and 83, respectively. Bilateral fittings were proportionally greater in audiometric groups b and c (20 and 18%, respectively) than groups a and d (9 and 12%, respectively). For the analysis of the archetypical patterns of benefit, the heterogeneous nature of this subset of the HSSS dataset – the breadth of hearing abilities as well as age and hearing aids represented – allows a robust stratification of the overall data, as well as the data within each audiometric groups, into benefit profiles (Eisenack et al., 2019).

### Data-driven pattern identification

The data-driven analysis used here was based on unsupervised and supervised learning and was divided into five main steps illustrated in Figure 1. The first step reduced the dimensionality of the data by applying factor analysis. The second step divided the processed dataset in four subsets of the data corresponding to the listeners belonging to four audiometric groups. The third step was the archetypal analysis which was used to identify the archetypal patterns of benefit. The listeners were then clustered in benefit profiles in the fourth step. Finally, the predictor importance of each of the items of the questionnaires was inferred from the fifth step.

#### I. Dimensionality reduction

Based on factor analysis (Cattell, 1988), the multi-dimensional dataset was reduced to four latent factors. The number of factors was selected by parallel analysis (Horn, 1965) with subsequent iterative resampling (K = 200). The factors were obtained using oblique Procrustes rotation as in (Akeroyd et al., 2014). The factor scores corresponding to each of the subjects, and obtained by Bartlett’s method, were used as the input of the archetypal analysis stage (III below).

#### II. Stratification

The listeners were divided into four groups corresponding to the audiometry-based auditory profiles a, b, c and d as suggested in Sanchez-Lopez et al. (2020). The binary rules used here are

a. Audiometric group-a: HL_HF_ < 50 dB HL, and HL_LF_ < 30 dBHL.
b. Audiometric group-b: HL_HF_ > 50 dB HL, and HL_LF_ < 30 dBHL.
c. Audiometric group-c: HL_HF_ > 50 dB HL, and HL_LF_ > 30 dBHL.
d. Audiometric group-d: HL_HF_ < 50 dB HL, and HL_LF_ > 30 dBHL.

#### III. Archetypal analysis

Matrix factorization was applied to the results of the dimensionality reduction step. A given observation was represented as a convex combination of the archetypal patterns (Cutler & Breiman, 1994). The analysis retrieves two matrices – the ‘pattern matrix’, which contained archetypal patterns represented in the data and the ‘subject matrix’, consisted of the weights corresponding to each pattern that resemble each of the observations. The specific implementation of the method used here was similar to Mørup & Hansen (2012). The identified patterns were ranked based on the degree of disability and handicap in each archetypal pattern of benefit and labeled based on rehabilitation needs as in clinical triage as APoB0, APoB1, APoB2, and APoB3, being APoB0 the optimal benefit pattern and APoB3 the suboptimal.

#### IV. Clustering

The distance between observations and the four archetypal patterns was estimated using the weights contained in the subject matrix. The criteria used here was the nearest archetype (Ragozini et al., 2017). Each observation was then assigned to a cluster based on their weights. While some observations had similar weights, no observation was equidistant from any two clusters. The “benefit profiles” were labeled according to the archetypal patterns of benefit (BP0, BP1, BP2, and BP3).

#### V. Importance estimation

Once the subjective data corresponding to each of the subjects were analyzed with unsupervised learning techniques, supervised learning was used for estimation the importance of the specific items of the dataset. A decision tree ensemble was trained with a subset of the data corresponding to the items of the SSQ and the HHQ and the identified clusters as the output. The ensemble was trained with 200 surrogated trees using curvilinear prediction. The importance was obtained by the permutation of out-of-bag features, which provides the minimum square error averaged for each tree over the standard deviation across the trees.

## Results

The present study sought to identify patterns of benefit associated with different audiometric groups. The data from the questionnaires SSQ and HHQ, collected in a heterogeneous group of hearing-aid users, was first explored using factor analysis. Then, the patterns of benefit were inferred by using archetypal analysis over the factor scores corresponding to four factors (speech understanding, spatial hearing, qualities of hearing and hearing handicap). The dataset was stratified into four subsets corresponding to the four audiometric groups. In addition, it was of interest to identify the priorities of hearing rehabilitation. This was done by evaluating the predictor importance of the individual items of the SSQ and HHQ. Once the participants were divided into clusters or *benefit profiles*, the results corresponding in each subdomain were explored with the purpose of applying the present findings in a personalized hearing rehabilitation strategy.

### Factor analysis

The parallel analysis revealed four factors as the optimal number of factors. The factor analysis was repeated in each of the stratified groups to confirm their similarity before further analysis. The loadings in each group were similar to the ones from the analysis of the entire dataset with small deviations commiserate with the changes in benefit patterns described below. Overall, the four factors corresponded to the three subdomains of the SSQ – SU, SP and QH – and hearing handicap (HH). These factors, taken together, explained a total of 50% of the variance.

The highest loading items in each of the three SSQ-based factors were similar to those of the validated short-form of the questionnaire, the SSQ12 (Noble et al., 2013). The SSQ12 is a twelve-item questionnaire with the same SU, SP and QH subdomains that was the result of an item selection process between three parties and based on different criteria, involving the scores reported in the previous factor analysis of the SSQ (Akeroyd et al., 2014). Given the familiarity with this version of the SSQ, and lack of need for another, similar version of the SSQ (cf. Demeester et al., 2012; Jensen et al. 2009; Moulin et al., 2019; von Gablenz et al., 2018), the remainder of the results are based on the SSQ12 as well as the HHQ.

### Data-driven analysis

Figure 2 shows the results of the data-driven analysis. The left panel reflects the archetypal patterns of benefit (APoB) with respect to the latent factors for each of the audiometric groups (a-d; lowercase to distinguish from the Sanchez-Lopez et al. (2020) auditory profiles). The analysis of the overall data is indicated by the dotted lines. The left panel reflects the “archetypal patterns” (APoB) with respect to the latent factors. The right panel represents the estimated importance of the individual items of the questionnaire. In the figure, only the three most important predictors in a given subdomain are shown for simplification.

**Figure 2.**
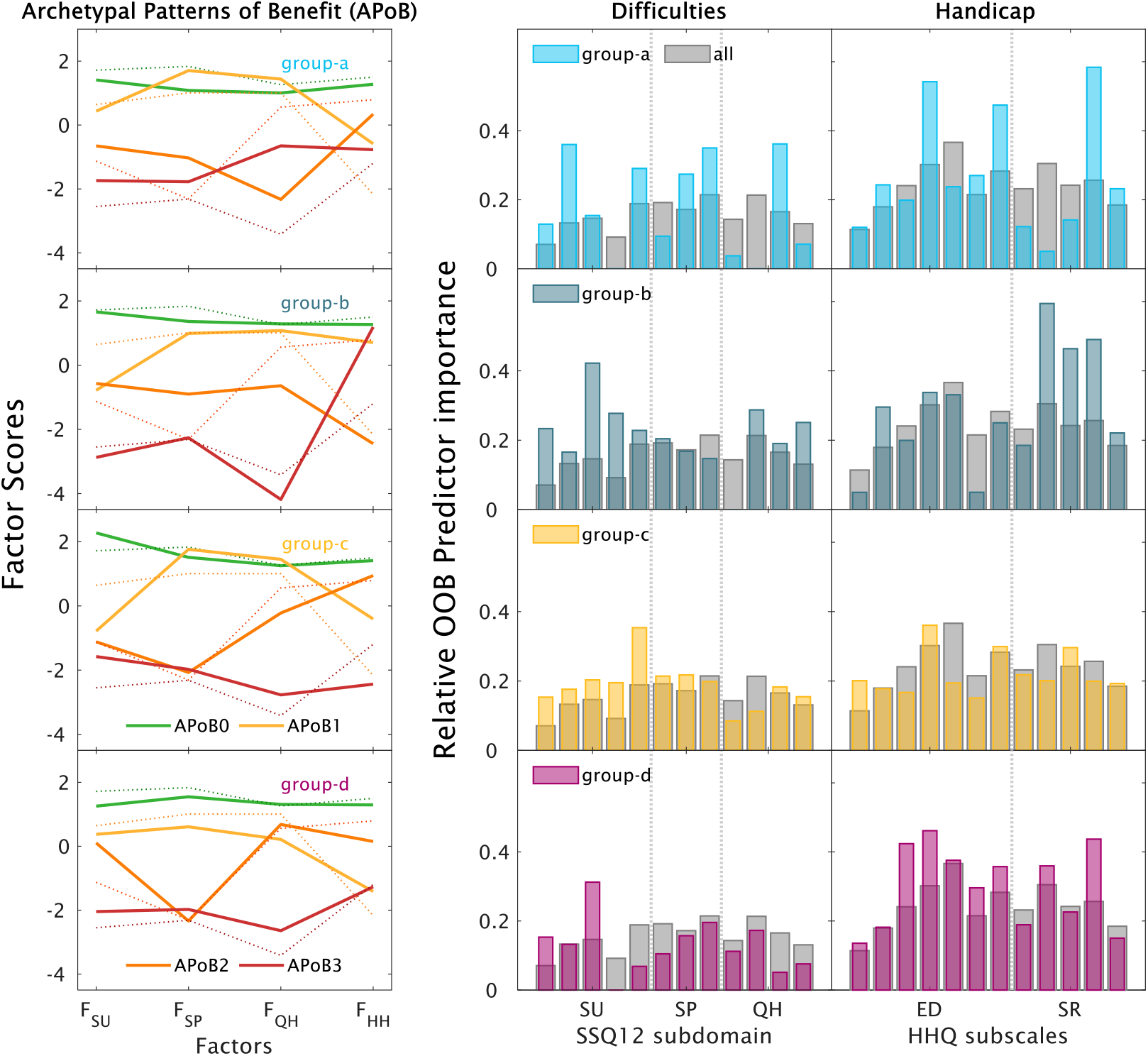
Data-driven analysis of subjective data stratified into audiometric groups. Each row corresponds to an audiometric group (a-d). Left panel: Archetypal patterns of benefit (APoB) resulting from step III of the method. Four patterns are ranked and labelled between optimal (APoB0; green) to suboptimal (APoB3; red). The analysis of the overall data is shown by the dotted lines. Right panel: Relative importance of the individual items estimated by the Out-of-the-bag permuted features delta error of a random forest. The twelve items of the SSQ12 are divided into three subdomains, speech understanding (SU), spatial hearing (SP), and qualities of hearing (QH), and the twelve items of the HHQ are divided into two subscales, emotional distress (ED) and social restrictions (SR). The grey bars show the results of using the same procedure on the entire dataset.

### Archetypal patterns of benefit

Figure 2 (left panel) shows the result of the archetypal analysis. The optimal profile (APoB0) showed a high score for all factors (green), which was similar for all of the audiometric groups, as well as for the entire data set (dotted lines). The near-optimal pattern (APoB1, yellow) was different for the different audiometric groups. For group-a and group-d, the pattern showed high scores for the difficulties-related factors (F_SU_, F_SP_, F_QH_) but a lower score for the handicap-related scores (F_HH_). In contrast, for group-b and group-c, APoB1 showed lower scores reflecting the difficulties in speech understanding (F_SU_). The four groups differed substantially in terms of the near-suboptimal pattern (APoB2, orange). For group-a, the lowest score corresponded to quality-related difficulties (F_QH_), whereas for group-b, the lowest score reflected an increased handicap. For group-c and group-d, the lowest score corresponded to difficulties with spatial hearing, while the handicap-related scores were higher than in APoB1. The suboptimal pattern (APoB3, red) showed the lowest scores for all factors in group-c. In group-d, APoB3 represented the lowest scores for SU, QH and HH but not for SP. However, for group-a the scores reflecting speech understanding and spatial hearing factors were lower than the scores reflecting qualities, while group-b showed higher scores in the items reflecting hearing handicap. The archetypal patterns of benefit corresponding to the analysis of the overall data (dotted lines) resembled, to a large extent, the patterns observed in group-d.

### Item importance estimation

The importance of the subdimensions in terms of difficulties and restrictions were estimated by the predictor importance of the individual items. The importance was considered here as indicative of priorities for hearing rehabilitation. Figure 2 (middle panel) shows the predictor importance corresponding to the items of the SSQ12. The predictor importance is shown for each of the audiometric subgroups (in color) and the overall data (in grey). The highest importance shown in the analysis of the overall data corresponded to the items related to emotional distress (ED), social restrictions (SR) and the spatial hearing (SP) difficulties. In contrast, the questions related to speech understanding (SU), and hearing qualities (QH) questions were found to be less important.

When analysing each audiometric group independently, the relative importance of the subdomains was substantially different than the one showed in the analysis of the entire dataset. Group-a showed higher importance than the overall group for SU in questions related to conversations with multiple talkers in quiet, SP difficulties in terms of distance, QH difficulties in the clarity of sounds and three HH questions on self-confidence and social restrictions. Group-b showed higher importance for SP difficulties related to speech-in-noise perception, QH difficulties related to listening effort and HH related to social restriction. Group-c showed similar importance for SP and HH as the overall data. The importance obtained for SU difficulties was higher than overall specifically for the ability to get the start of the sentences during conversational turn-taking. Group-d showed higher importance for speech-in-noise difficulties compared to the overall data, and the highest importance generally for the HH subdomain.

All audiometric groups also showed less importance than overall for some items (i.e., when the coloured bars are much lower than the grey overall scores in the middle and right panels of Figure 2). This importance, though, should only be interepreted as these items not being useful in the prediction of the benefit profiles. For group-a there was relatively less importance for noisy group conversations (SU), identifying music (QH) and avoiding social situations (HH). For group-d, noisy group conversations (SU) was also relatively less important. For group-b, there was relatively less importance for distinguishing multiple sound sources (QH) and feeling self-conscious (HH). For group-c, the only relatively less important item hearing-related nervousness/discomfort (HH).

Overall, the stratified approach for the analysis of priorities for hearing rehabilitation revealed different patterns of importance for the different audiometric groups of listeners.

### Benefit profiles and audiometric groups

Once the participants have been stratified into audiometric groups (stage II of the method summarized in Figure 1) and divided into benefit profiles (stege IV), the data corresponding to the SSQ12 items and the HHQ is presented across the subdomains of the difficulties and handicaps in Figure 3. Each row corresponds to an audiometric group (a-d) and each column corresponds to a subdomain (SU, SP, QH and HH). The boxplots corresponding to the four benefit profiles are presented in each subplot. Furthermore, the significant differences between profiles, the result of pairwise comparisons using Bonferroni’s correction, are presented using brackets and asterisks.

**Figure 3:**
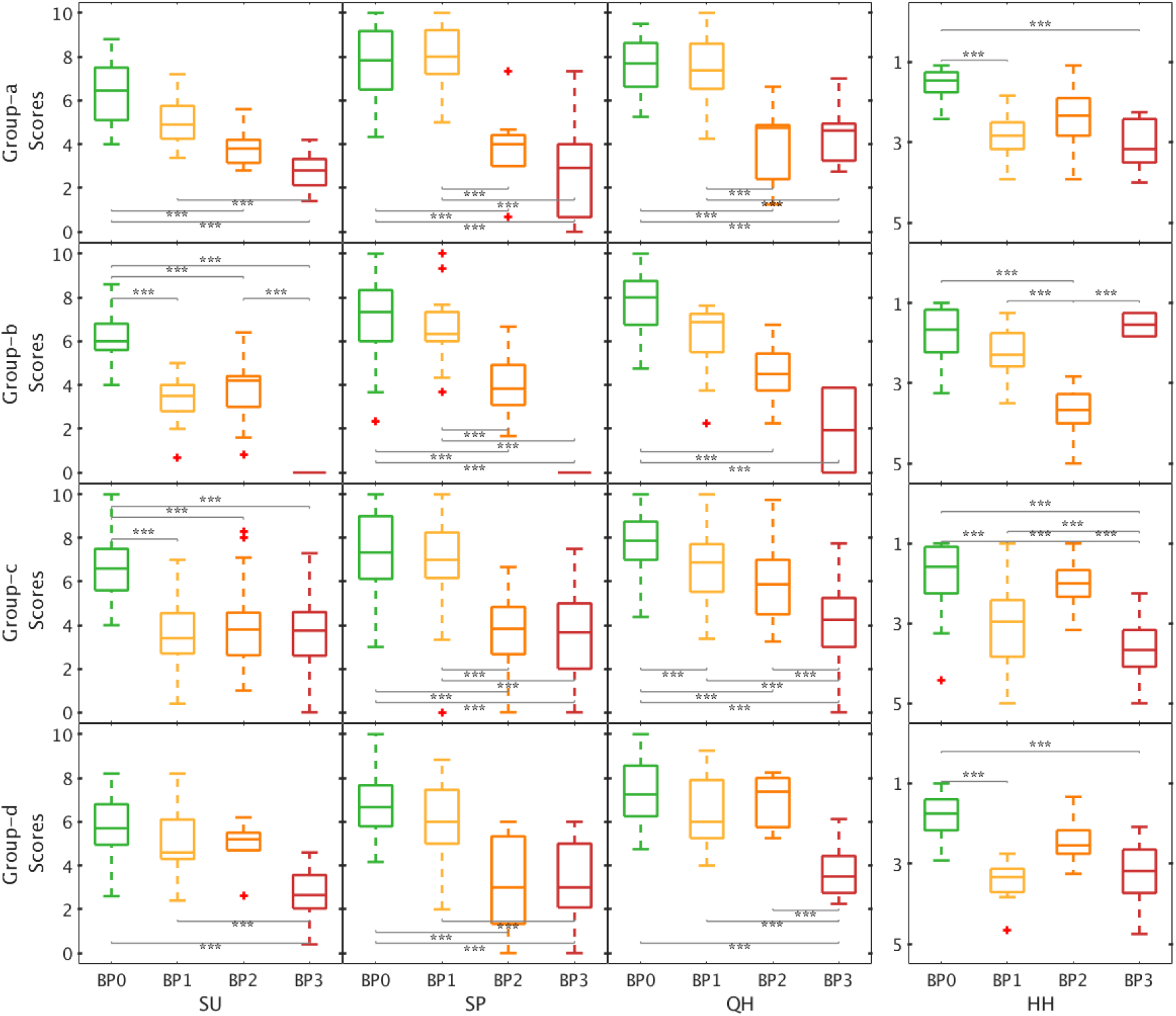
Average item scores for each benefit profiles. The results are shown for each audiometric group (columns) and for each subdomain disability and handicap subdomains (columns). SU: speech understanding; SP: spatial hearing; QH: qualities of hearing, and HH: hearing handicap. The y-axis for the HH results (far right column) is reversed for comparison to the other results, as HH scores (lower = less handicap) represent the inverse of SU, SP and QH responses (lower = more difficulty). The lines with (***) refer to significant differences p < 0.0001 with Bonferroni corrections.

The optimal profile (BP0) was similar in all subgroups with scores above 6 for the three subdomains of the SSQ12 and below 2 for the HHQ. The suboptimal profile (BP3) showed more variability across audiometric groups; median BP3 scores were less than 4 for the SSQ12 and greater than 3 for the HHQ except for group-b which showed HHQ much lower handicap (i.e., a median score near 1).

In group-a, benefit profiles BP2 and BP3 were significant different than the optimal profile (BP0) for speech, spatial and qualities of hearing. However, only BP1 and BP3 showed significantly worse scores than BP0 for the hearing handicaps (right panel). Furthermore, BP1 and BP2 were significantly different for spatial hearing and qualities. In group-b, the BP1-3 were significantly lower than BP0 for speech understanding. For spatial hearing and qualities of hearing, only BP2 and BP3 were significantly lower than BP0. In group-c, BP1-3 all showed significantly lower scores than BP0 for speech and qualities of hearing. In terms of handicap, BP1 and BP3 showed significantly lower scores than BP0, whereas BP2 was not significantly lower than BP0. Finally, for group-d, only the BP3 was significantly lower than the optimal benefit profile for speech understanding and qualities of hearing. However, BP1 was significantly lower for hearing handicap.

The present results showed that the benefit profiles BP1 and BP2 showed significant differences with the optimal (BP0) and suboptimal (BP3) profiles. The patterns observed in each row suggested that each audiometric group might have different needs for improvements to reach similar scores as BP0.

### Significant differences between audiometric groups

Overall, the results presented in Figure 3 showed differences in benefit profiles across the subdomains of hearing difficulties and hearing handicaps. Figure 4 shows the same data but focused on the differences among the audiometric groups.

**Figure 4:**
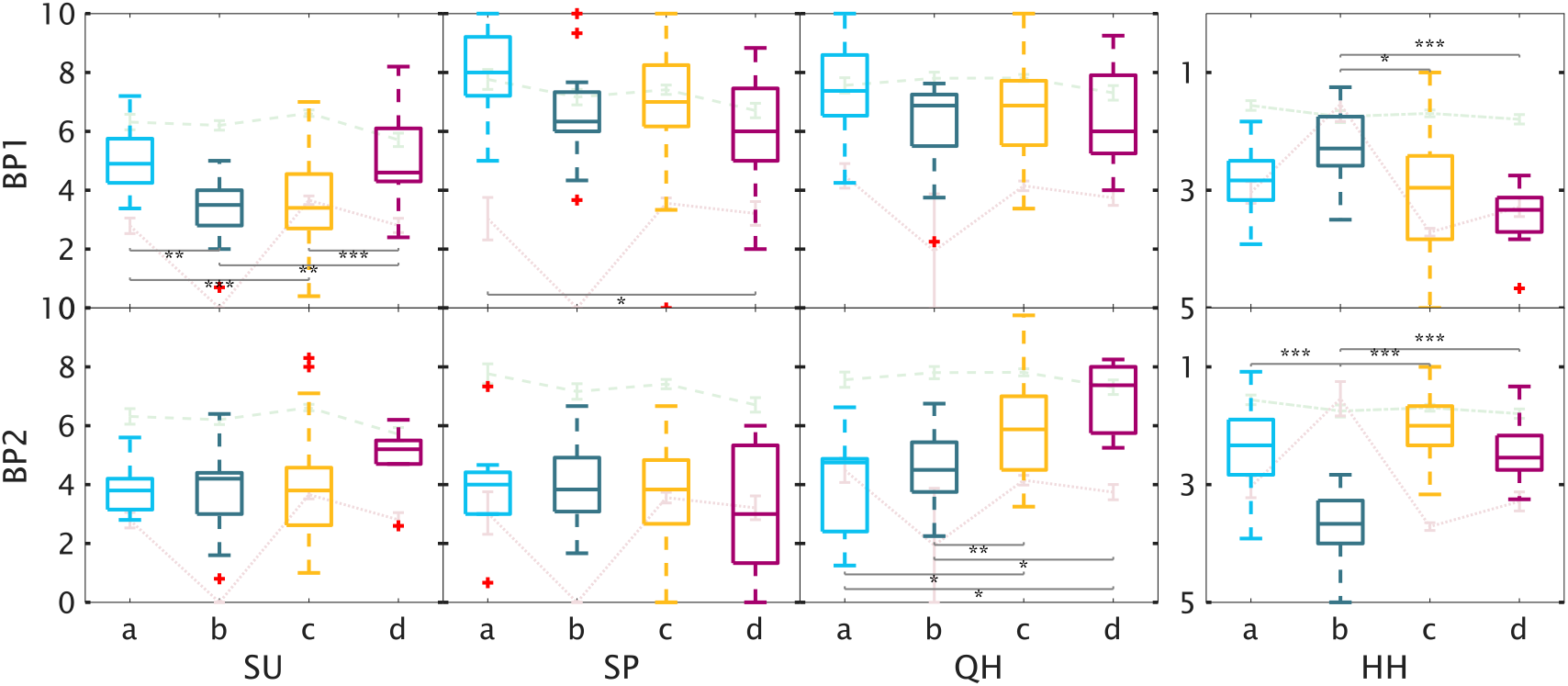
Average item scores for each audiometric groups. The results are shown for the benefit profiles BP1 (top row) and BP2 (bottom row) and for each subdomain disability and handicap subdomains (columns). SU: speech understanding; SP: spatial hearing; QH: qualities of hearing, and HH: hearing handicap. The scores of the HH subdomain are shown in a reverse y-axis since 1 corresponds to “good” and 5 to “poor” in the HHQ questionnaire. The lines with (***) refer to significant differences *p < 0*.*0001 (**)* to *p < 0*.*001* and (*) *p < 0*.*01* with Bonferroni corrections.

The top row shows the BP1 for the four audiometric groups and across the subdomains. The green dashed line reflects the optimal profile and the red dotted line the suboptimal profile. The four audiometric groups showed significant differences in speech understanding and hearing handicap. The participants in group-b and group-c showed lower scores for SU while group-a and group-d showed scores closer to the optimal benefit profile. In terms of hearing handicap, group-c and group-d showed significantly higher (better) scores than group-b and inline with BP0. The bottom row shows the near-suboptimal profile (BP2). The scores for SU and SP were not significantly different across the four auditometric groups whereas the scores for qualities of hearing were significantly lower for group-a and group-b. In terms of hearing handicap, group-b showed significantly higher (poorer) scores than the other audiometric groups.

## Discussion

### Audiometric groups and auditory profiles

The analysis of the self-reported hearing difficulties and handicaps identified patterns of benefit and priorities for continued post-fitting rehabilitation. The participants were stratified into audiometric groups based on the audiometric distinctions (i.e., LF and HF thresholds) in the auditory profiles from Sanchez-Lopez et al. (2020). Those auditory profiles were the result of a data-driven analysis of multidimensional data that included measures of audibility, loudness perception, speech perception, binaural processing abilities and spectro-temporal resolution. Despite being derived from the previous auditory profiles A-D, the audiometric groups a-d used in the present study to stratify the participants cannot be considered equivalent to those profiles. There are notable similarities and discrepancies between the objective hearing deficits in the previous study’s auditory profiles and the subjective difficulties and handicaps associated with the current study’s audiometric groups and their benefit profiles.

The current study’s audiometric group-a listeners showed, on average, a high importance of speech communication in quiet, spatial perception difficulties and a high priority for rehabilitating hearing handicaps (Figure 2, first row), whereas in Sanchez-Lopez et al. (2020), auditory profile A listeners showed a low degree of perceptual deficits and a close-to-normal speech recognition. Audiometric group-b listeners had a single priority for rehabilitation, difficulties in speech understanding (see BP1 in Figures 3 and 4), consistent with the reduced speech-in-noise perception performance in Profile B listeners. Audiometric group-c listeners showed priorities for rehabilitating difficulties in speech-in-noise perception and handicaps (as shown BP1 in Figure 4), moderately consistent with Profile C listeners who showed a high degree of perceptual deficits. Finally, the audiometric group-d listeners showed a priority for rehabilitating hearing handicaps followed by spatial hearing difficulties (BP1 and BP2 in Figure 4), whereas Profile D listeners showed near-normal suprathreshold perception except for their abnormal loudness perception. In summary, the one consistency in the objective and subjective outcomes of the auditory profiles and current audiometric groups, respectively, is in their difficulties with speech-in-noise.

The auditory deficits in the audiometric groups discussed above were associated with the residual difficulties observed in the near-optimal benefit pattern (BP1). The patterns of benefit revealed that these residual difficulties and handicaps were in many cases independent, and minor difficulties do not always imply minor handicaps. This can be observed in audiometric groups a and d, where the near-optimal pattern of benefit corresponded to high scores in the SSQ12 items but low scores in the HHQ items. This is in agreement with Noble et al. (2013), who concluded that SSQ12 should be accompanied by the HHQ to provide a complete evaluation of the level of hearing disability and handicap.

### Interpretation of the patterns of benefit

The data-driven analysis revealed four benefit profiles, labelled BP0, BP1, BP2, BP3. BP0, the “optimal” benefit profile, corresponds to patients with relatively minimal difficulties and handicaps who do not require additional intervention and may only need periodic follow-up visits (e.g., once per year). BP0 was characterized by scores above 6 in the SSQ12 and below 2 in the HHQ, similar to previous findings (Noble, Naylor, Bhullar, & Akeroyd, 2012). The HCP should ensure that this optimal result does not change by evaluating the intervention periodically. BP1, the “near-optimal” benefit profile, corresponds to patients who would require some adjustments or instructions in regular follow-up visits to improve the treatment. BP1 was characterized by a significant reduction of only one or two subdomain scores (i.e., HH for groups a and d, SU for group b, and SU and QH for group c), but not in others (e.g., SP), as shown in Figure 3. The HCP should be aware of the limitations and allocate time to perform these improvements successfully. BP2, the “near-suboptimal” profile, corresponds to an intervention that requires major improvements. BP2 was characterized by a reduction of the scores in at least two subdomains compared to the scores of BP0 (e.g., looking at Figure 3 first and third rows, group-a and group-c showed lower scores for SU, SP and QH but not for HH). Patients who show this pattern might reflect problems that require substantial adjustments and additional intervention through structured sessions focused on alleviating different difficulties and handicaps and/or providing assistive listening devices and training. BP3, the “suboptimal” profile, corresponds to low benefit. BP3 was characterized by the relatively greatest difficulties and handicaps. This suggests that the initial intervention should be reconsidered. In this case, the HCP should evaluate the possibilities of changing the device (e.g. considering a bone-anchored device if there are conductive loss indications) as well as evaluate the need for a multi-disciplinary approach (e.g., intensive auditory training, clinical psychological interventions).

### Personalized hearing rehabilitation

The present study shows the potential of data-driven analysis for providing insights for improving the efficiency and efficacy of the hearing healthcare services. The KDD approach applied in this study aims at providing new knowledge by applying suitable methods to relevant datasets and focused on the interpretability of the results to obtain new knowledge. However, the perspectives discussed in this section should be taken as an outlook of the possibilities of this approach and not as a direct application of the findings from this study.

Personalized hearing healthcare could be introduced in the hearing care clinic in the form of stratified audiology. Since different patterns of difficulties and handicaps were observed in different audiometric groups, the patients can be first stratified into meaningful groups associated with specific hearing deficits and rehabilitation priorities to minimize the confounds of sensory hearing deficits. In the present study this was initially done by using pure-tone audiometry. Pure-tone audiometry can be complemented by speech-in-noise perception tests, the assessment of loudness perception, binaural processing abilities or spectro-temporal modulation sensitivity to better characterize the individual’s hearing deficits (Sanchez-Lopez et al., 2019). When such characterizations are not possible, the results of the current study show that stratification based solely on limited audiometric information can provide additional personalisation when coupled with self-reported outcomes.

The common healthcare goal is an “optimal” intervention which would minimize the activity limitations and participation restrictions by providing the most suitable rehabilitation for each individual. In such a personalized hearing healthcare approach, the outcome measures can be used to identify the benefit profile of the patient, a profile associated with specific and important difficulties and restrictions result from the data-driven approach. The use of, for example, SSQ12 and HHQ in a follow-up visit after an acclimatization period would be useful for guiding the need for further testing and intervention which can be more time-consuming but can provide a meaningful input for the intervention. Furthermore, the present results suggest that an evaluation of the intervention based on SSQ12+HHQ can lead the decision-making, but the intervention required in each audiometric group might need a different pathway. Alternatively, open-response questionnaires such as the Client-Oriented Scale of improvement (COSI; Dillon et al., 1997) could help set individual priorities based on the important situations. COSI is typically used, though, for setting goals and expectations, guiding the hearing-aid selection and to quantify the improvements after the intervention, but not planning the intervention (Dillon & So, 2000). Open-response questionnaires can be very useful for identifying hearing difficulties; a future archetypal analysis focused on social situations may provide greater insight. Nevertheless, participation restrictions have been previously shown to be better evaluated using structured questionnaires such as HHQ (Stephens et al., 2000) to help the decisions for further intervention. Based on the benefit profile derived from these outcomes, a personalized rehabilitation program can be offered to the patient where the intervention can then be focused on systematically overcoming specific hearing difficulties or handicaps with the help of the present stratification. The goal would be to obtain an optimal profile (close to BP0) at the end of the rehabilitation program.

The intervention is often prioritized in terms of sensory management, perceptual training and counselling in a “holistic approach” (Boothroyd, 2007). In contrast, an initial post-fitting assessment with items from the SSQ12 and HHQ based on the present findings, as well as tempering expectations (Whitmer et al., 2016), could effectively guide further rehabilitation beyond the initial intervention (i.e., after the hearing-aid fitting). For example, the present study revealed that some difficulties were more important than others for a given audiometric group. Speech-in-noise was important for audiometric groups a and c but speech-in-speech was more important for groups b and d. The former would lead to a device adjustment (e.g., increasing the scale of noise reduction or making the “noisy/restaurant” setting the default), whereas the latter would lead to some rehabilitative training on listening strategies (e.g., how best to position yourself in conversation). These insights can also be useful for recreating the important situations in a clinical environment and then evaluate the improvement by means of aided performance tests. In cases with handicap priorities in their profile, the participation restrictions can be evaluated using measures out of the clinic that can capture the user positive and negative experiences in daily life (Lund et al. 2020). One last important factor that can be part of this personalized hearing healthcare is to gather information about the auditory ecology of the patient (Gatehouse et al., 2003) which could be captured by the active responses of the patient (ecological momentary assessments; e.g., Holube et al., 2020; Smeds et al., 2020) or their hearing devices (Christensen et al., 2019).

#### Limitations of the study

The main limitation of the current study is that the data used in this analysis was collected between 2002-2011. Hearing-aid technology has evolved substantially since then. The ratio of bilateral fittings is also currently higher than it was when this data was collected. The lower ratio of bilateral fittings represents a potential confound when interpreting the spatial hearing difficulties in this group. Applying this archetypal analysis to a more recent dataset might show different results, as residual difficulties and handicaps may have diminished, although one would not expect the patterns or profiles to substantially change.

A related limitation is the lack of additional information about the hearing-aid fitting provided in the dataset. The data does not contain, for example, real-ear measurements or details about the advanced features of the hearing aids nor the acoustic coupling. Furthermore, there is no information related to any burdens or issues associated with the use of hearing aids, which can be important for establishing viable measures of hearing-aid benefit (Whitmer et al. 2016).

A limitation of the proposed personalization is the risk of using group profiling as a unique characterization instead of individual profiling. The benefit profiles simplifying the complex problem of evaluating each individual’s hearing-aid benefit. The patients’ responses are the result of a retrospective judgement that often reflects their average hearing experience but can also be influenced by specific events or individual factors (Saunders, Chisolm, & Abrams, 2005; Wu, Stangl, Zhang, & Bentler, 2015). While this approach can simplify the personalization process, the unique characterization of each individual’s benefit and rehabilitation needs often requires the expertise of the HCP in asking additional questions about the user experiences and the openness and consistency of the patient explaining their residual difficulties and restrictions. Therefore, the flexibility and expertise a well-trained HCP provides to the hearing rehabilitation is something that the envisioned personalized strategy should not affect detrimentally, but work in favour of a better service.

## Conclusions

A data-driven approach applied to self-reported residual difficulty and handicap outcomes revealed four different benefit profiles -- optimal, near-optimal, near-suboptimal and suboptimal -- for each of four clinical subpopulations based on the audiometric differences in Sanchez-Lopez et al. (2020). The optimal benefit profiles for each audiometric group set the difficulty and handicap minima that can be used to determine the scale and type of post-fitting rehabilitation necessary to improve outcomes for those with near-optimal to suboptimal profiles. Using short questionnaires, this simplified personalisation scheme promotes a form of personalised care that can be implemented clinically or remotely. The application of these patterns of benefit and the use of stratification could improve clinical interventions of hearing loss as well as the efficiency and quality of service in hearing care clinics.

## Data Availability

The data used in the present study is only available under request.

## Acknowledgments

The authors thank A. Ahrens and O. Cañete for their comments in an earlier version of the manuscript. The authors acknowledge the valuable feedback from the members of the Hearing Sciences Scottish Section during the realization of the present study.

